# Post-Acute COVID-19 Effects on Diagnostic Conversion Rates And Standardized Cognitive and Motor Test Scores in a Longitudinal Study of Independent, Community-Recruited Elderly Subjects

**DOI:** 10.1101/2025.10.20.25338408

**Authors:** Nathaniel Dunckley, Nan Zhang, Charles H. Adler, Holly A. Shill, Shyamal Mehta, Erika Driver-Dunckley, Christine M. Belden, Alireza Atri, Parichita Choudhury, Angela Kuramoto, Geidy E. Serrano, Thomas G. Beach

## Abstract

There is considerable concern about the long-term consequences of COVID-19 infection, generally referred to as post-acute sequelae, including declines in cognitive and motor abilities. The degree to which COVID-19 contributes additional burden to normal aging trajectories remains unclear. Additionally, the impact of COVID-19 on subjects under study for age-related neurological diseases is a potential confounder that needs definition. This study investigated whether having had a COVID-19 illness was associated with a differential decline in cognitive or motor function in older adults, utilizing data from the Arizona Study of Aging and Neurodegenerative Disorders (AZSAND) and Brain and Body Donation Program (BBDP), a longitudinal clinicopathological study based in metropolitan Phoenix, Arizona. Subjects were included if they 1) had completed a questionnaire about their experience with COVID-19 illness 2) were classified as cognitively normal at pre-pandemic diagnostic cognitive consensus conferences and 3) had one or more subsequent diagnostic conferences between July 1, 2020 and August 30, 2025. All subjects had serial standardized research-dedicated clinical evaluations including the Montreal Cognitive Assessment (MoCA) and the Unified Parkinson’s Disease Rating Scale (UPDRS). Specific objectives were to compare, between those who reported having had or not having had COVID-19, rates of conversion to cognitive impairment or dementia as well as pre-pandemic and final MoCA and UPDRS motor scores. A total of 100 subjects self-reported having had COVID-19 while 71 denied having had it. Their related acute illness severity was generally mild, with only 10% having had hospital treatment and none having required ventilator support. Post-acute symptoms were also mild; only 1 subject reported having “long Covid”. Both cognitive and motor performance, as measured by MoCA and UPDRS part 3 scores, declined slightly (not statistically significant) over the study period. Conversion of cognitive diagnosis from normal to impaired or dementia occurred in 26% of those having had COVID-19 and in 32% of those not having had COVID-19; the difference was not significant. All subjects diagnosed with PD at the start of the study were also diagnosed with PD at the end of the study; no subjects converted from not having to having probable PD. Logistic regression analysis indicated that subjects’ report of having had COVID-19 was not significantly associated with conversion to cognitive impairment or dementia while greater age, male sex, possession of one or more apolipoprotein E-ε4 alleles, and a diagnosis of probable PD all conferred a significantly greater likelihood of conversion. The results suggest that having a mild COVID-19 illness is not associated with greater declines on cognitive or motor screening tests than would be expected from age-related changes alone. Limitations of this analysis include small sample sizes, potential misclassification of COVID-19 status, and reliance on relatively crude clinical metrics that may miss significant functional changes. Additionally, we were unable to separately assess for varying COVID-19 severity dependent on whether or not the subject had been vaccinated, the number and type of vaccinations, and the particular SARS-CoV-2 variants that were circulating at the time of illness.

## INTRODUCTION

The COVID-19 pandemic has sparked widespread concern about the long-term consequences of COVID-19 infection on neurological health, generally referred to as post-acute neurological sequelae. Alongside respiratory and systemic complications, many patients have reported cognitive, motor, and sensory symptoms, including memory difficulties, fatigue, movement abnormalities, and anosmia [1–3]. These concerns have given rise to a growing amount of research examining whether or not COVID-19 may accelerate neurodegenerative processes.

A number of other viral infections have previously been linked to neurological damage or increased risk for neurodegeneration. For example, herpesviruses have been implicated in Alzheimer’s disease [4], while influenza has been associated with cognitive impairment and inflammatory damage in the brain [5]. Recent postmortem studies of COVID-19, including work by Serrano et al. [6], have similarly reported molecular and inflammatory changes in brain regions such as the olfactory bulb and amygdala, even when direct viral detection is limited. These findings have led researchers to explore whether viral infections can trigger or amplify neuropathological processes, particularly in vulnerable populations. Alongside these findings, it is reasonable to ask whether COVID-19 may similarly contribute to long-term neurological decline.

The relationship between COVID-19 infection and aging-related neurological decline remains poorly defined. Some large-scale retrospective studies and electronic health record analyses have suggested increased risk for dementia and cognitive impairment following infection [7], while others have raised questions about confounding variables, such as pre-existing conditions and pandemic-related stressors [8]. Furthermore, many studies do not incorporate long-term, pre-pandemic baseline data for direct comparison, which limits their ability to distinguish between normal age-related cognitive changes and COVID-19 associated changes [12].

This issue is particularly relevant for older adults, who already face elevated risk for neurodegenerative disorders. The degree to which COVID-19 contributes additional burden to their normal aging trajectories or preclinical disease remains unclear. Distinguishing self-reported symptoms from measurable changes in cognitive and motor function is essential to developing an accurate understanding of the virus’s long-term impact. Additionally, the differential impact of COVID-19 on subjects under study for age-related neurological diseases is a potential confounder that needs definition in order to avoid misinterpretation of data.

The Arizona Study of Aging and Neurodegenerative Disorders (AZSAND) and Brain and Body Donation Program (BBDP) [10] is a longitudinal clinicopathological study and as such provides valuable data to assess these questions in a well-characterized aging cohort. With decades of longitudinal data on cognitive and motor, combined with detailed demographic and neuropathological information, this study enables comprehensive tracking of functional change over time.

By comparing pre- and post-pandemic changes in standardized cognitive and motor research assessments this study investigated whether or not having had a COVID-19 illness was associated with a differential decline in cognitive or motor function in older adults.

## METHODS

### Subject Selection

Subjects were selected by database searches of the AZSAND/BBDP, a longitudinal clinicopathological study based in greater Phoenix, Arizona (10). A subset of subjects were co-enrolled in the National Institute on Aging-funded Arizona Alzheimer’s Disease Research Center. Search criteria specified that subjects 1) had completed a questionnaire about their experience with COVID-19 illness (see Supplementary Methods), including stating whether or not they had ever had COVID-19; 2) were classified as cognitively normal at diagnostic cognitive consensus conferences between November 1, 2018 and August 30, 2020; and 3) had one or more subsequent diagnostic conferences between July 1, 2020 and August 30, 2025. Subsequent diagnostic conferences after the first noted whether or not subjects had “converted” to several forms of mild cognitive impairment (MCI), dementia or non-specific cognitive impairment. Subjects who converted were only included in the study if the cognitive conference first noting the conversion occurred after their first completed COVID-19 questionnaire. Subjects who never developed cognitive impairment were only included if, at conferences between January 1, 2024 and August 30, 2025, they were still classified as cognitively normal and denied having had COVID-19 on their final questionnaire administered over the same time period.

### Subject Characterization

All subjects had serial standardized research-dedicated clinical evaluations, done by teams of nurses, medical assistants, behavioral neurologists, movement disorder neurologists, neuropsychologists and psychometrists using standardized assessment batteries [10], including, for most subjects, the National Alzheimer’s Coordinating Center (NACC) Uniform Data Set [21], the Montreal Cognitive Assessment (MoCA) [22] and the Unified Parkinson’s Disease Rating Scale (UPDRS) [23]. Following subject testing, consensus conferences were held to establish diagnoses with respect to cognitive status and presence or absence of movement abnormalities including the presence or absence of probable Parkinson’s disease.

### Statistical Analysis

Demographic information was analyzed using unpaired, 2-way t-tests and Fisher Exact tests. Scores on the MoCA and Part 3 of the UPDRS were analyzed using one-way analysis of variance (ANOVA) with post-hoc Tukey paired significance testing. The relationship between having had COVID-19 and conversion to impairment or dementia was examined using logistic regression, with covariables including age, sex, years of education and presence or absence of a probable Parkinson’s disease (PD) diagnosis. All variables were converted to dichotomous values for ease of comparing odds ratios. Age of 82 or older, male sex, and education years of 16 or greater were all assigned a value of 1.

## RESULTS

### COVID-19 Diagnosis, Severity and Symptoms

A total of 171 subjects met inclusion criteria; 100 reported having had COVID-19 (COVID group) while 71 reported not having had it. Table 1 gives demographic data for the two groups. Of those who reported having had COVID-19, 83 reported that on at least one occasion this had been on the basis of a positive nasal swab test; the chemical nature of the test done was not addressed by the questionnaire and the number of separate COVID-19 illnesses or infections sustained by each subject was not determined. The groups were not significantly different in terms of age, sex, education, possession of the apolipoprotein E ε4 allele, and proportion having a diagnosis of probable PD. All subjects diagnosed with PD at the start of the study were also diagnosed with PD at the end of the study; no subjects converted from not having to having probable PD.

**Table 1.**
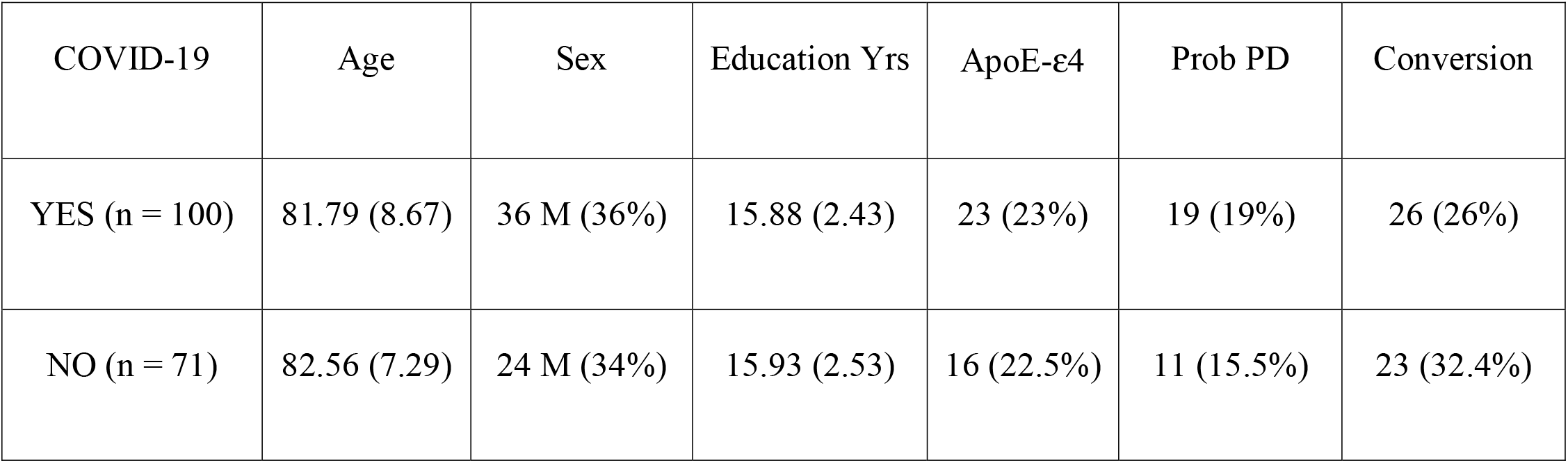
Demographic characteristics of subjects reporting having had or not having had COVID-19. The age of subjects given here is their age at study end, August 30, 2025. Conversion refers to the number and percentage of subjects that converted from cognitively normal to cognitive impairment or dementia. Means and standard deviations are shown for age and years of education. Groups were not significantly different in terms of age, sex, education, possession of the apolipoprotein ε4 allele, proportion having a diagnosis of probable Parkinson’s disease (PD) or proportion converting from cognitively normal to cognitively impaired or dementia.

| COVID-19 | Age | Sex | Education Yrs | ApoE-ε4 | Prob PD | Conversion |
| --- | --- | --- | --- | --- | --- | --- |
| YES (n = 100) | 81.79 (8.67) | 36 M (36%) | 15.88 (2.43) | 23 (23%) | 19 (19%) | 26 (26%) |
| NO (n = 71) | 82.56 (7.29) | 24 M (34%) | 15.93 (2.53) | 16 (22.5%) | 11 (15.5%) | 23 (32.4%) |

Of 99 subjects responding to the question about whether they went to a hospital because of a COVID-19 diagnosis, 10 answered affirmatively while 89 denied this; hospital stays were limited to one or two days. No subjects had been put on a ventilator while 4 received oxygen. Nine received antibiotics, 3 received corticosteroids, 1 received remdesivir. Symptoms included 30 with cough, 21 with fatigue, 19 with sore throat, 19 with muscle aches, 13 with fever and/or chills, 12 with headache, 12 with runny nose, 11 with shortness of breath, 9 with nausea and/or vomiting and/or diarrhea, 7 with loss of taste and/or smell, and 6 with wheezing. Only 4 subjects reported having had difficulty concentrating, memory problems or “brain fog”. None affirmed having had a transient ischemic attack, stroke, heart attack or kidney failure. Generally, less than 2 or 3 subjects, for each symptom, reported that they were severe. Only 1 subject reported having “long COVID” or “long-hauler syndrome” while 36 denied having it. Only 2 of the 100 subjects reporting having had COVID-19 reported not having had a vaccination for it. All 71 of the subjects not reporting having had COVID-19 reported having been vaccinated at least once.

### Cognitive and Motor Changes During Study Period

Over the study period, conversion of cognitive diagnosis from normal to impaired or dementia occurred in 26% of those that reported having had COVID-19 and in 32% of those reporting not ever having had COVID-19; the difference in proportion was not significant.

There were no significant differences for declines in either MoCA or UPDRS part 3 scores (Table 2) between those reporting having had COVID-19 and those that denied having had COVID-19.

**Table 2.** Comparison of pre- and post-pandemic MoCA and UPDRS motor scores in subjects reporting having had or not having had COVID-19. Means and standard deviations are shown. Analysis of variance indicates a significant difference (p = 0.02) across groups for MoCA scores; paired significance testing, however, indicates a significant difference (p = 0.019) only for the comparison of pre-pandemic and post-pandemic scores in those reporting having had COVID-19 versus those not reporting having had COVID-19, respectively (asterisks). There is no significant difference across groups for UPDRS scores.

| COVID-19 | MoCA<br>Study Beginning | MoCA<br>Study End | UPDRS<br>Study Beginning | UPDRS<br>Study End |
| --- | --- | --- | --- | --- |
| YES | 26.56 (2.08) n = 48* | 25.96 (2.57) n = 48 | 2.95 (4.37) n = 60 | 4.79 (7.63) n = 60 |
| NO | 26.37 (2.03) n = 40 | 24.8 (4.16) n = 40* | 2.40 (5.67) n = 42 | 4.90 (6.27) n = 42 |

Logistic regression analysis (Table 3) was conducted to determine the relationship of self-reported COVID-19 on the likelihood for conversion from cognitively normal to cognitively impaired or dementia, when also considering subject age, sex, years of education, possession of an apolipoprotein E ε-4 allele and having had a diagnosis of probable PD. Subjects’ report of having had COVID-19 was not significantly associated with conversion while greater age, male sex, possession of one or more ApoE-ε4 alleles, and a diagnosis of probable PD, all conferred a significantly greater likelihood of conversion.

**Table 3.** Logistic regression analysis of relationship of reported COVID-19 on likelihood for conversion from cognitively normal to cognitively impaired or dementia, when also considering subject age, sex, years of education, possession of an apolipoprotein E ε-4 allele and having had a diagnosis of probable Parkinson’s disease (PD). All variables were converted to dichotomous values for ease of comparing odds ratios. Age of 82 or older, male sex, and education years of 16 or greater were all assigned a value of 1. Subjects’ report of having had COVID-19 was not significantly associated with conversion while age, sex, ApoE - ε4 and a diagnosis of probable PD all had significant associations with conversion. * for variables significantly associated with conversion.

| Variable | Odds Ratio (95% CI) | Probability | Beta Coefficient | Standard Error |
| --- | --- | --- | --- | --- |
| Reported COVID-19 | 0.71 (0.335-1.52) | 0.38 | -0.338 | 0.386 |
| Age* | 4.67 (1.91-11.46) | 0.0007 | 1.54 | 0.457 |
| Male Sex* | 3.76 (1.70-8.30) | 0.0010 | 1.328 | 0.404 |
| Education Years | 1.42 (0.62-3.25) | 0.405 | 0.352 | 0.422 |
| ApoE - ε4* | 3.16 (1.32-7.55) | 0.0095 | 1.151 | 0.444 |
| Probable PD* | 2.865 (1.00-8.20) | 0.050 | 1.052 | 0.536 |

## DISCUSSION

This study investigated whether COVID-19 infection is associated, over several years after the acute stage, with measurable declines in motor and cognitive function in older adults who were initially classified as cognitively unimpaired. The results showed that there was not a significantly greater rate of conversion to cognitive impairment or dementia in the COVID-19-positive group. and between-group comparisons of MoCA and UPDRS motor scores did not yield statistically significant differences.

These findings are generally in agreement with the prior literature (Table 4), with most studies finding little or no objective evidence of an increased rate of cognitive decline in those that had a clinical COVID-19 infection. When differences between COVID-19 and non-COVID-19 were found, they were mostly limited to subjective symptoms.

**Table 4.** Summary of relevant publications.

| Study | Findings |
| --- | --- |
| Arbula et al., 2024 [11] | Case-control study of post-COVID patients with cognitive complaints showed attentional deficits and higher anxiety/depression. There was a notable mismatch between subjective complaints and objective impairments, suggesting psycho-affective contributions. |
| Caselli R.J et al., 2023 [12] | Longitudinal cohorts with pre and post COVID infection neuropsychological testing found little evidence of accelerated within-person cognitive decline after COVID-19. Memory, executive, language, and behavioral measures remained stable. |
| Cataldo S.A. et al., 2024 [13] | Long-COVID clinic cohort reported prominent cognitive complaints, yet objective testing showed little to no impairment. MRI analysis revealed significant decreased volume in multiple key brain areas, such as the cerebellum, in long-COVID patients. |
| Hampshire A. et al., 2024 [9] | Large community sample showed objectively measurable cognitive deficits after COVID. Resolved covid cases had small deficits, while unresolved persistent symptoms had larger deficits. Cognitive impacts were strongest for memory, reasoning, and executive tasks |
| Holland J. et al., 2023 [14] | Irish cohort study linked self-reported immune changes to subjective cognitive complaints in post-COVID syndrome. Relationship persisted even after accounting for depression, fatigue, sex, BMI, and education. Findings suggest immune dysregulation contributes to cognitive symptoms beyond mood effects. |
| Leitner M. et al., 2024 [15] | Survey of 416 long-COVID patients found high prevalence of fatigue, memory problems, brain fog, and dyspnea. Symptoms fluctuated and worsened after activity. Nearly half reported professional/financial losses, reduced working hours, and diminished stress tolerance. Vaccination shortened acute illness duration and delayed long-COVID onset. Women reported more symptoms than men. |
| Levine K.S. et al., 2023 [16] | Genetic biobank study (FinnGen & UK Biobank) linked prior viral infections to elevated neurodegenerative disease risk. COVID not the main focus, but influenza, pneumonia, and encephalitis showed strong associations with Alzheimer's, Parkinson's, ALS, and dementia. Provides indirect evidence that viral illnesses (including SARS-CoV-2) may influence long-term neurodegenerative risk. |
| Panagea E. et al., 2025 [17] | Review summarized evidence that long-COVID is associated with persistent cognitive impairments ("brain fog," memory loss, executive dysfunction). Emphasized |
|  | heterogeneity: some patients show objective deficits, while others primarily report subjective complaints. Highlights need for standardized assessment tools and longitudinal designs to resolve inconsistencies. |
| Schild A-K. et al., 2024 [18] | Longitudinal follow-up of non-hospitalized PCS patients. Reported impairments in memory, concentration, and attention consistent with SARS/MERS data. Found mild to moderate impairment (16–26%), persisting up to 12 months. Anxiety/depression increased over time. Authors note trajectories vary and highlight need for standardized cognitive rehabilitation approaches. |
| Sekendiz Z. et al., 2023 [19] | Compared pre- and post-COVID cognitive functioning. Found minimal objective changes across global cognition, but many patients reported persistent subjective complaints. Suggests possible mismatch between subjective experience and objective measures, similar to other PCS studies . |
| Szewczyk W. et al., 2024 [20] | Focused on subjective cognitive decline in long-COVID. Found high prevalence of perceived deficits, particularly memory and attention. Many participants linked symptoms to fatigue, anxiety, and depression. Emphasized that subjective decline often exceeded objective impairment, suggesting affective contributions. |

Our study is one of very few to examine objective motor and movement outcomes after COVID-19. We found no significant differences in UPDRS motor score declines on the basis of having had or not having had COVID-19. Two studies that looked at the post-acute experience of COVID-19 in patients with PD and other pre-existing neurological disorders [25,26] found that those with COVID-19 had greater risks of dyspnea, fatigue, falls, brain fog and inflammatory myopathies compared to those without COVID-19. Levodopa dose adjustment was higher post-infection in those that had COVID-19. In our study, there were no subjects that converted to PD and for those that had PD at the start of the pandemic, the MoCA and UPDRS motor scores for those that had COVID-19 were not significantly different, at study beginning or at study end, from those that did not have it (results not shown); however, these comparisons were of limited power due to very small sample sizes for those that had the scores (n = 5 per group for MoCA score; n = 2 for UPDS scores).

It is important to realize that, in contrast to these apparently minor post-acute effects of COVID-19 on our studied population, acute SARS-CoV-2 infection was common and had fatal consequences for many of our subjects [6,27]. Of 2,099 questionnaires filled out by 824 subjects over the study period, 357 subjects (43%) reported having had COVID-19 on at least one questionnaire. Furthermore, of 310 subjects who died between January 1, 2020 and May 22, 2025, and had a postmortem nasopharyngeal swab analyzed for SARS-CoV-2 with an FDA-approved clinical PCR test, 60 were positive (19%), and, of the 46 who tested positive and had a complete autopsy, 25 also had pneumonia, sufficient as a COVID-19 related cause of death. This suggests that having a perimortem SARS-CoV-2 infection had fatal consequences in about 50% of our cases.

Aside from the significance of post-COVID-19 sequelae for the general population, the pandemic introduced a new variable that may affect clinical diagnostic conversion and progression rates and so underscores the importance of using longitudinal, pre-pandemic baselines to assess the true impact of COVID-19 on neurological aging.

Limitations of this analysis include missing data, modest sample sizes for some comparisons, potential misclassification of self-reported COVID-19 status, relatively mild acute COVID-19 illnesses, and reliance on relatively crude clinical metrics that may have missed significant functional changes.

The error rate for self-reporting COVID-19 is uncertain. A literature review found only one study that assessed this; self-report had sensitivity and specificity of 90.5% and 96.5%, respectively, when compared to a SARS-CoV-2 PCR test administered in hospital an average of 16 weeks earlier [28]. Our study subjects were considerably older, however, and therefore might be expected to have less accurate recall of their health history.

Other limitations were that we were unable to separately assess for varying COVID-19 severity dependent on whether or not the subject had been vaccinated, the number and type of vaccinations, and the particular SARS-CoV-2 variants that were circulating at the time of illness. Vaccinations for SARS-CoV-2 first became FDA-approved in the US in December 2020, and although we are missing first vaccination dates for most of our subjects, of those that listed dates, all were in the first few months of 2021. As more than 90% of subjects’ first questionnaires were completed after March 2021 and more than 80% of the first reports of having had Covid were in questionnaires from January 2022 or later, this suggests that the great majority of subjects reporting having had COVID-19 had already had a vaccination prior to their first illness. Therefore our study may underestimate the severity of post-acute COVID-19 sequelae as the illness is known to be more severe in unvaccinated individuals.

## Supporting information

Questionnaire example

## Data Availability

All data produced in the present work are contained in the manuscript.

