## Supplementary material for "Post-Acute COVID-19 Effects on Diagnostic Conversion Rates And Standardized Cognitive and Motor Test Scores in a Longitudinal Study of Independent, Community-Recruited Elderly Subjects": Questionnaire example

IRB Approved at the
Study Level
Dec 07, 2021

### Brain and Body Donation Program Covid-19 Medical History Research Questionnaire

**Version 5, 11/18/2021; Thomas G. Beach, MD, PhD**

Introduce to participant as: "This questionnaire is to gather information about your experience with Covid-19, also known as Coronavirus, also known as SARS or SARS- CoV-2".

**Date of Questionnaire Administration/Interviewer Name:** ______________________________________________

**BBDP Participant Name/DOB/Donor ID:** ______________________________________________________________

**BBDP Study Partner Name/DOB (month /year only):** _____________________________________________________

**Person Completing Questionnaire (circle all that apply):** Participant  Study Partner or Relative  BBDP Personnel

**Questionnaire Supplemented by Medical Records Review (circle):** Yes  No  Deferred

**Have you had Covid-19, or an illness caused by coronavirus (circle):** Yes  No  Not Sure

**Has a healthcare provider ever told you that you had Covid-19 (circle)** Yes  No  Not Sure
If Yes, please provide healthcare provider's name and phone number: __________________________________

**Have you ever been tested by nose swab for Covid-19 (circle):** Yes  No  or Not Sure
If Yes, what was the approximate date of the test?  Exact Date_____________or Month/Year ______________
If Yes, what was the test result (circle):  Positive  Negative  Indeterminate  or Not Sure

**Have you ever had a blood test for Covid-19 (circle):** Yes  No  or Not Sure
If Yes, what was the approximate date of the test?  Exact Date_____________or Month/Year _______________
If Yes, what was the test result (circle):  Positive  Negative  Indeterminate  or Not Sure

**Where was Covid-19 testing done (circle all that apply):** Doctor's Office Urgent Care  Medical Lab  Hospital Drive-Up

**Did you ever go to a hospital because of Covid-19?** Yes No or Not Sure
If Yes, what hospital: ______________________

If Yes, Exact Dates________________or Month/Year ______________

If Yes, what doctor/provider took care of you there: ____________________________________ or Not Sure

**Where in the hospital were you (circle all that apply):** Emergency Inpatient ICU Covid-19 area or Not Sure

**What breathing treatment did you have (circle all that apply):** None Oxygen Ventilator ECMO or Not Sure

**What medicines were you treated with (circle all that apply):** or Not Sure

Azithromycin Other antibiotics Dexamethasone Inhaled Corticosteroids (flovent, symbicort, Advair)

Oral corticosteroids (prednisone) Remdesivir Convalescent Serum Hydroxychloroquine/Choroquine

Anti-coagulants (blood thinners) Monoclonal antibody Pfizer Covid pill (Paxlovid)

Merck Covid pill (molnupiravir)

**How long were you in the hospital (circle):** Few Days Week or More Month or More or Not Sure

**Did participant die as a result of Covid-19 illness?** Yes No

**Have you been having, or having with increased frequency or severity (relative to previous years) of any of these symptoms over the past year? (circle all that apply):**

Fever Chills Shakes Muscle Aches Runny Nose Sore Throat Loss of Taste Loss of Smell

Headache Fatigue Difficulty Concentrating Cough Wheezing Shortness of Breath/Difficulty Breathing Chest Pain Nausea Vomiting Abdominal Pain Diarrhea Stroke Transient Ischemic Attack (TIA)
Myocardial Infarction (Heart Attack) Kidney Failure

**Have you ever been vaccinated or immunized for Covid-19 or coronavirus? (circle)** Yes No or Not Sure

**If yes, what type of vaccine was it? (circle one of the choices below):**

Pfizer Moderna Astra-Zeneca Johnson & Johnson Janssen Novavax or Not Sure

If Yes, what was the approximate date of the vaccination? Exact Date ___________or Month/Year __________

If the vaccination required a second dose, give date here. Exact Date ___________or Month/Year __________

**Have you ever had a booster vaccination for Covid-19 or coronavirus? (circle)** Yes No or Not Sure

**If yes, what type of vaccine was it? (circle one of the choices below):**

Pfizer Moderna Astra-Zeneca Johnson & Johnson Janssen Novavax or Not Sure

If Yes, what was the approximate date of the vaccination? Exact Date ___________or Month/Year __________
